# Duration of SARS-CoV-2 Sero-Positivity in a Large Longitudinal Sero-Surveillance Cohort: The COVID-19 Community Research Partnership

**DOI:** 10.1101/2021.01.27.21250615

**Authors:** David Herrington, The COVID-19 Community Research Partnership Study Group

## Abstract

**Background:** Estimating population prevalence and incidence of prior SARS-CoV-2 infection is essential to formulate public health recommendations concerning the COVID-19 pandemic. However, interpreting estimates based on sero-surveillance requires an understanding of the duration of elevated antibodies following SARS-CoV-2 infection, especially in the large number of people with pauci-symptomatic or asymptomatic disease.

**Methods:** We examined >30,000 serology assays for SARS-CoV-2 specific IgG and IgM assays acquired longitudinally in 11,468 adults between April and November 2020 in the COVID-19 Community Research Partnership.

**Findings:** Among participants with serologic evidence for infection but few or no symptoms or clinical disease, roughly 50% sero-reverted in 30 days of their initial positive test. Sero-reversion occurred more quickly for IgM than IgG and for antibodies targeting nucleocapsid protein compared with spike proteins, but was not associated with age, sex, race/ethnicity, or healthcare worker status.

**Interpretation:** The short duration of antibody response suggests that the true population prevalence of prior SARS-CoV-2 infection may be significantly higher than presumed based on earlier sero-surveillance studies. The impact of the large number of minimally symptomatic COVID-19 cases with only a brief antibody response on population immunity remains to be determined.

**Funding:** This publication is supported by the CARES Act, of the U.S. Department of Health and Human Services (HHS) as part of a financial assistance award totaling $20,000,000. The contents are those of the author(s) and do not necessarily represent the official views of, nor an endorsement, by HHS, or the U.S. Government.

**Research in context**

Evidence before this study
Previous longitudinal studies of the humoral response to SARS-CoV-2 suggest that people with less severe disease have a more rapid decline of SARS-CoV-2 specific antibodies than people with severe disease. However, these data come from small laboratory-based investigations or studies of convenience samples identified based on symptomatic disease.

Added value of this study
This study provides extensive longitudinal serologic follow-up in a large number of people with serologic evidence of prior infection who had little or no symptoms based on active daily symptom surveillance.

Implications of the available evidence
The data indicate that serologic evidence of prior infection in minimally symptomatic people is fleeting, suggesting that cross-sectional sero-surveys have under-estimated the true prevalence of prior infection in populations. The data highlight the challenge of determining transmission dynamics and long-term immunity in asymptomatic cases which likely represents an even larger fraction of all cases of prior SARS-CoV-2 infection than previously presumed.

## Introduction

Determining the proportion of the population previously infected with SARS-CoV-2 and how this rate has changed over time is essential to understand the pandemic and recommendations for clinical preparedness, physical distancing, targeting of vaccines, and resumption of economic activities. Unfortunately, tests for viral antigens or RNA in symptomatic or high risk individuals are inadequate for this purpose because of the transient nature of viral shedding.

Sero-surveillance, especially when deployed in large, population-based samples is thought to provide more accurate estimates of the prevalence of prior SARS-CoV-2 infection. Indeed, several sero-surveillance studies have highlighted the fact that a significant proportion of previously infected people are pauci- or completely asymptomatic and therefore likely missed by clinically motivated testing.^1,2^ These data illustrate the importance of using testing strategies that include minimally and asymptomatic cases when estimating community transmission.

However, sero-surveillance for SARS-CoV-2 infection has important limitations. In addition to the well described issues related to the sensitivity and specificity of different serologic assays ^3,4^ there is also uncertainty about the expected duration of elevated antibodies following SARS-CoV-2 infection. Understanding the dynamics of the humoral response is important as it has a direct impact on completeness of ascertainment when using sero-surveillance to determine population prevalence. The durability of the humoral response may also provide clues concerning the degree of immune activation following primary infections and the likelihood of subsequent long-term immunity in individuals and in the population. Preliminary evidence from small clinical studies suggests that minimally symptomatic infections often have an attenuated antibody response;^3,5-10^ however, more data are needed from large population samples with more detailed information on symptoms to complement the data from these intensive laboratory-based investigations.

Accordingly, we examined more than 30,000 longitudinally acquired serology test results from more than 11,461 adults enrolled in the COVID-19 Community Research Partnership - a population-based COVID-19 syndromic and sero-surveillance study based in two large healthcare systems in central North Carolina. The overwhelming majority of participants had few or no symptoms of COVID-19 even though more than 10% had serologic evidence of infection. Thus, this study provides a unique opportunity to examine the durability of antibody responses in a population-based survey including the large and critically important portion of the population with asymptomatic or pauci-symptomatic infection.

## Methods

Beginning on April 16^th^, 2020 potential participants 18 years and older identified in the Wake Forest Baptist Health (WFBH) and the Atrium Health (AH) systems were invited to participate through email, internal communications, websites, and social and general media. After providing informed consent, participants were asked to record daily symptoms (e.g., fever, cough, shortness of breath, etc.) related to COVID-19^11^ using a web-based Patient Monitoring System application (Oracle Corporation, Redwood Shores, California). A subset of participants (serology cohort) was also selected for longitudinal sero-surveillance based on their age, race, and gender to reflect the distribution of these demographics in their county of residence^12^, with oversampling of certain high-risk groups (health care workers and minorities).

Participants selected for sero-surveillance were mailed kits for in-home testing of finger-prick capillary blood. Initially participants received a Syntron Bioresearch Inc. lateral flow assay (LFA) to test for IgM and IgG antibodies to the SARS-CoV-2 nucleocapsid antigens (n=13,752 assays). In-home LFA results were recorded and interpreted via a smartphone application with central review (Scanwell Health, Inc. © 2020). A subset of participants received two 20 µL volumetric absorptive microsamplers (Mitra®, Neoteryx) for blood collection that were analyzed centrally using the same Syntron LFA (n=4,313 assays). In July, 2020 the Syntron assay became unavailable after which participants received the EUA approved Innovita Biological Technology Co. lateral flow assay (LFA) to test for IgM and IgG antibodies to the SARS-CoV-2 spike and nucleocapsid antigens (n=16,868 assays). Both assays were validated at the Frederick National Laboratory for Cancer Research (FNLCR) by the National Cancer Institute (NCI) using a panel of antibody-positive samples from patients with PCR confirmed SARS-CoV-2 infection or pre-pandemic controls (Panel 2); Syntron: (antibody: sensitivity/specificity); IgM: 93.3%/97.5%; IgG:73.3%/100%; IgM or IgG:96.7%/97.5%), Innovita: (antibody: sensitivity/specificity); IgM:93.3%/98.8%; IgG: 93.3%/98.8%; IgM or IgG:100%/97.5%.^13^ Additional validation of these tests in a point-of-care format produced similar results.

The number and cadence of tests performed by each participant was influenced by the rolling enrollment into the cohort over time (earlier enrollees had more time for serial testing), as well as several factors related to the pandemic including interruptions in supply chains and test kit availability, shipping delays to and from the participants, and variability in the rate participants completed in-home tests or returned specimens for in-lab testing. Thus, estimates of sero-reversion in this report are derived from samples of the entire seroconversion cohort over a range of times following an initial positive test rather than assessment of the entire cohort at precisely timed intervals. The number and cadence of testing was similar among those with at least one positive test during follow-up and those that remained negative (Supplemental Figure 1).

Conventional parametric measures of central tendency and variance were used unless the distribution suggested that other approaches (e.g. Poisson confidence intervals) were more suitable. Logistic regression was used to estimate the relative odds of seroconversion as a function of symptom prevalence (JMP Ver. 15.0, SAS Institute). Multivariable Weibull^14^ and semi-parametric Cox proportional hazard^15^ models for interval-censored data were used to estimate the survival curve of time to sero-reversion controlling for age, self-reported, race/ethnicity, healthcare worker status, and enrolling healthcare system. The Wald test based on bootstrap standard errors was used for significance testing of the parameter estimates. (R package icenReg, v 3.63^16^).

Role of the funding source: This work was supported by a grant from the State of North Carolina funded by the CARES Act, of the U.S. Department of Health and Human Services (HHS). The sponsor had no role in the developing the study design; in the collection, analysis, and interpretation of data; in the writing of the report; or in the decision to submit the paper for publication.

## Results

Between April 16^th^ and Jan. 4^th^, 2020 11,468 participants aged 18-94 yrs. completed a total of 30,620 serologic tests for IgM or IgG antibodies to SARS-CoV-2 antigens (tests/participant: range: 1-8; mean+/-95%CI_(Poisson)_ 2.67+/-2.64-2.70; Table 1, Figure 1). During the period of observation 1,172 people had at least one positive test for either IgG or IgM (crude sero-prevalence = 10.2%). Active daily symptom monitoring beginning at enrollment confirmed that COVID-19 symptoms were uncommon in this seropositive cohort. A COVID-like illness (defined as fever plus cough or shortness of breath for two out of three consecutive days) in the month prior to serology testing was associated with a positive result (OR=11.4, p<0.0001); but was reported in only 4% of seropositive participants. Similarly, two of three consecutive days of fever, sore throat, cough, shortness of breath, chest pain, muscle pain, nausea, diarrhea, headache, or anosmia were individually associated with subsequent seroconversion when present (all p≤0.0004), but were infrequently reported (symptom prevalence range: 1%-17%).

**Figure 1.**
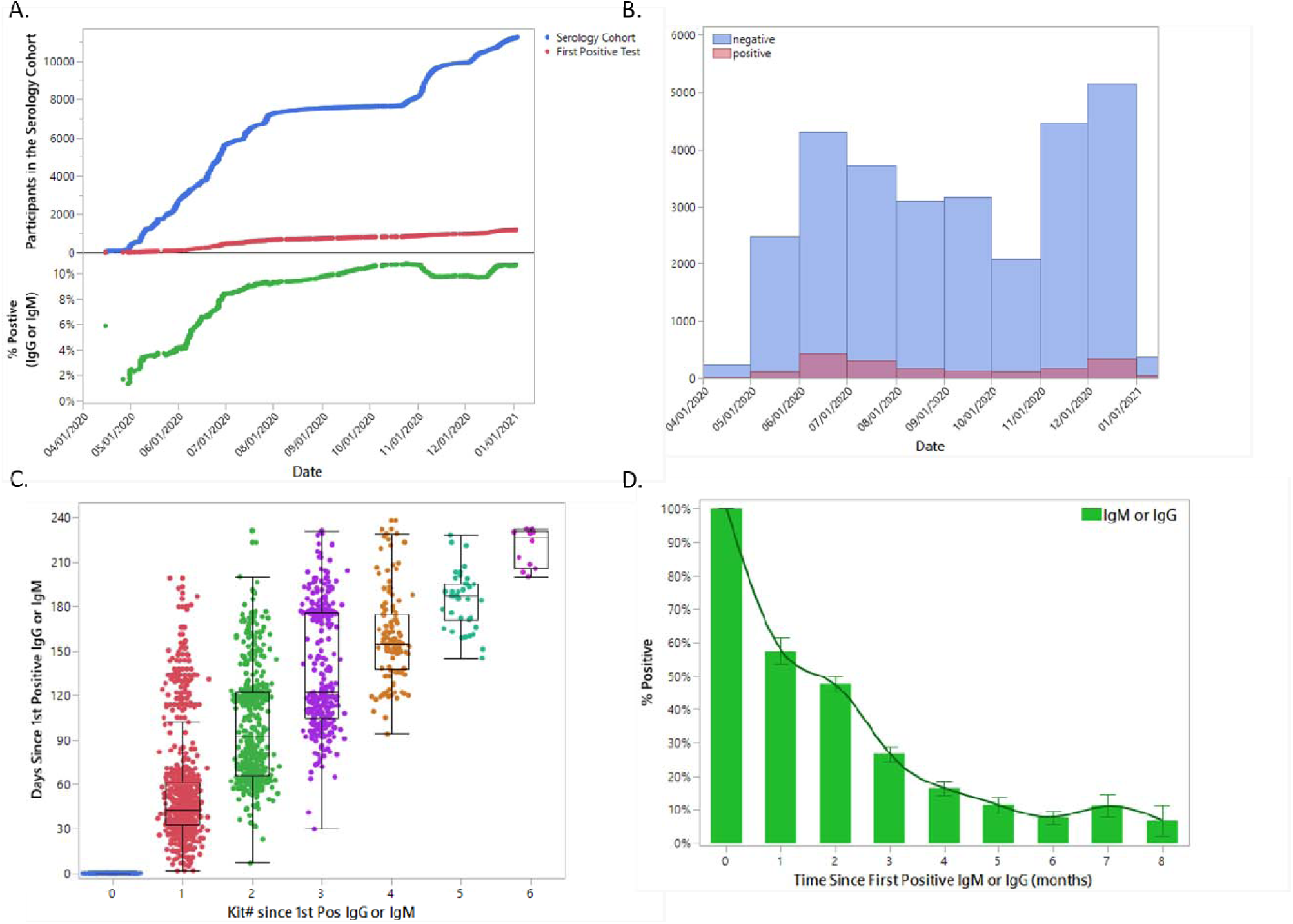
Distribution of Enrollment of Participants and Serology Tests as a Function of Time. Panel A. Number of participants enrolled, and number and percent of participants who seroconverted from April 4th 2020 to Jan. 9th 2021. Panel B. Number of positive and negative tests from April 4th 2020 to Jan. 9th 2021. Panel C. Distribution of longitudinal tests following an initial positive result. Panel D. Percent of positive tests as a function of time following initial seroconversion.

**Table 1.** Participants in the Sero-Survey.

|  | **Serology Cohort**<br>(n = 11,468) |  |  | **Seropositive Sub-Cohort**<br>(n = 1,172) |  |  |
| --- | --- | --- | --- | --- | --- | --- |
| **Total Number of Tests** | 30,620 |  |  | 3,856* |  |  |
| **Age (yrs)** | *n* | *%* | *tests/person(mean)* | *n* | *%* | *tests/person (mean)* |
| 1. <30 | 1003 | 8.7% | 2.4 | 99 | 8.5% | 3.0 |
| 2. 30-39 | 2357 | 20.6% | 2.8 | 258 | 22.0% | 3.3 |
| 3. 40-49 | 2420 | 21.1% | 2.8 | 244 | 20.8% | 3.4 |
| 4. 50-59 | 2486 | 21.7% | 2.7 | 258 | 22.0% | 3.4 |
| 5. 60-69 | 2079 | 18.1% | 2.6 | 210 | 17.9% | 3.3 |
| 6. $\geq 70$ | 1123 | 9.8% | 2.5 | 103 | 8.8% | 3.0 |
| **Sex** |  |  |  |  |  |  |
| F | 7085 | 61.8% | 2.7 | 719 | 61.4% | 3.3 |
| M | 4383 | 38.2% | 2.7 | 453 | 38.7% | 3.3 |
| **Race/Ethnicity** |  |  |  |  |  |  |
| Black or African American | 622 | 5.4% | 2.1 | 71 | 6.1% | 2.2 |
| Hispanic or Latino | 351 | 3.1% | 2.2 | 42 | 3.6% | 2.4 |
| Other | 554 | 4.8% | 2.4 | 55 | 4.7% | 2.9 |
| White (not Hispanic/Latino) | 9941 | 86.7% | 2.7 | 1004 | 85.7% | 3.4 |
| **Healthcare Worker** |  |  |  |  |  |  |
| N | 6949 | 60.6% | 2.4 | 629 | 53.7% | 3.1 |
| Y | 4519 | 39.4% | 3.1 | 543 | 46.3% | 3.5 |
| **Healthcare System** |  |  |  |  |  |  |
| Atrium Health | 2589 | 22.6% | 2.5 | 298 | 25.4% | 2.9 |
| Wake Forest Baptist Health | 8879 | 77.4% | 2.7 | 874 | 74.6% | 3.4 |

\* including first positive and all subsequent tests

A small number of participants (n=56) reported a clinical diagnosis of COVID-19 prior to enrollment which was confirmed with their initial serology test. Another 13 participants developed symptomatic COVID-19 requiring hospitalization during follow-up. Collectively, these cases of clinically significant COVID-19 represent 6% of the seropositive cohort.

Of the 1,172 people with at least one positive test for either IgM or IgG, 770 participants had 1-6 additional tests over the following eight months (mean interval between tests = 47.8 days, Figure 1C). Among the 148 participants who completed their next test within 30 days only 85/148 (57%) remained positive for IgG or IgM (Table 2). The percent of positive tests from the seropositive cohort continued to decline to <10% over the next five months. A similar early decline in sero-positivity was observed when examining results for the IgM or the IgG assays individually.

**Table 2.** Test Results as a Function of Time Following an Initial Positive Result.

**Seropositive Cohort**
|  | Baseline* | Month 1 |  | Month 2 |  | Month 3 |  | Month 4 |  | Month 5 |  | Month 6 |  | > 6 Months |  |
| --- | --- | --- | --- | --- | --- | --- | --- | --- | --- | --- | --- | --- | --- | --- | --- |
| IgM | n | n | % | n | % | n | % | n | % | n | % | n | % | n | % |
| negative | 0 | 59 | 48.8% | 257 | 61.8% | 255 | 78.7% | 227 | 88.0% | 155 | 93.4% | 168 | 92.3% | 99 | 93.4% |
| positive | 973 | 62 | 51.2% | 159 | 38.2% | 69 | 21.3% | 31 | 12.0% | 11 | 6.6% | 14 | 7.7% | 7 | 6.6% |
| IgG |  |  |  |  |  |  |  |  |  |  |  |  |  |  |  |
| negative | 0 | 23 | 41.1% | 73 | 38.6% | 57 | 57.6% | 79 | 71.2% | 34 | 75.6% | 38 | 95.0% | 17 | 70.8% |
| positive | 532 | 33 | 58.9% | 116 | 61.4% | 42 | 42.4% | 32 | 28.8% | 11 | 24.4% | 2 | 5.0% | 7 | 29.2% |
| IgG_or_IgM |  |  |  |  |  |  |  |  |  |  |  |  |  |  |  |
| negative | 0 | 63 | 42.6% | 255 | 52.4% | 264 | 73.3% | 254 | 83.6% | 163 | 88.6% | 181 | 92.3% | 107 | 89.9% |
| positive | 1172 | 85 | 57.4% | 232 | 47.6% | 96 | 26.7% | 50 | 16.4% | 21 | 11.4% | 15 | 7.7% | 12 | 10.1% |
| IgG_and_IgM |  |  |  |  |  |  |  |  |  |  |  |  |  |  |  |
| negative | 0 | 19 | 65.5% | 74 | 64.4% | 44 | 71.0% | 52 | 81.3% | 23 | 85.2% | 25 | 96.2% | 8 | 72.7% |
| positive | 330 | 10 | 34.5% | 41 | 35.7% | 18 | 29.0% | 12 | 18.8% | 4 | 14.8% | 1 | 3.9% | 3 | 27.3% |
| **Subset with Initial Positive IgG and IgM** |  |  |  |  |  |  |  |  |  |  |  |  |  |  |  |
| IgG and IgM |  |  |  |  |  |  |  |  |  |  |  |  |  |  |  |
| negative | 0 | 19 | 65.5% | 74 | 64.4% | 44 | 71.0% | 52 | 81.3% | 23 | 85.2% | 25 | 96.2% | 8 | 72.7% |
| positive | 330 | 10 | 34.5% | 41 | 35.7% | 18 | 29.0% | 12 | 18.8% | 4 | 14.8% | 1 | 3.9% | 3 | 27.3% |
| IgG or IgM |  |  |  |  |  |  |  |  |  |  |  |  |  |  |  |
| negative | 0 | 13 | 44.8% | 44 | 38.3% | 33 | 53.2% | 38 | 59.4% | 17 | 63.0% | 22 | 84.6% | 6 | 54.6% |
| positive | 330 | 16 | 55.2% | 71 | 61.7% | 29 | 46.8% | 26 | 40.6% | 10 | 37.0% | 4 | 15.4% | 5 | 45.5% |
| **Subset with Negative Test $\leq 60$ Days Prior** |  |  |  |  |  |  |  |  |  |  |  |  |  |  |  |
| IgG_or_IgM |  |  |  |  |  |  |  |  |  |  |  |  |  |  |  |
| negative | 0 | 38 | 52.1% | 90 | 65.7% | 58 | 69.1% | 48 | 72.7% | 47 | 79.7% | 36 | 85.7% | 31 | 83.8% |
| positive | 371 | 35 | 48.0% | 47 | 34.3% | 26 | 31.0% | 18 | 27.3% | 12 | 20.3% | 6 | 14.3% | 6 | 16.2% |

\* defined as the first positive test

Some test results were likely false positives, making it difficult to know what portion of the early decline in test positivity was due to true sero-reversion versus simple correction of an original false positive result. To minimize the effect of false positives, we examined data from the smaller number of participants whose first positive test was positive for both IgG and IgM (specificity = 100% for both Syntron and Innovita based on NCI validation panels). Similar to the overall results, relatively few of these participants who were tested again in the first 30 days remained positive for both IgG and IgM (35%). Even when counting either IgG or IgM in the subsequent tests, the sero-positive rate was only 55% in the first 30 days following the initial positive test. In the second month following the initial positive test the test positive rate rose slightly to 62% but then steadily declined over the ensuing four months (Table 2).

For participants whose first test after enrollment was positive it is impossible to know how much time had passed since their primary infection. Therefore, we restricted the analysis to the 371 people whose first positive test was preceded by a negative test ≤60 days prior (mean, 95%CI = 38.8, 37.6-40.0 days). As in the full cohort, the test positive rate declined to less than 50% within 30 days and exhibited a steady decline to <15% over the ensuing five months (Table 2).

Based on analysis of the interval censored data, the estimated time to 50% sero-reversion for IgM or IgG was 35.7 days (Figure 2A). The rate of sero-reversion was not associated with age, sex, race/ethnicity, healthcare worker status or site of enrollment. The estimated time to sero-reversion was significantly faster in participants who were pauci- or asymptomatic compared with those with clinically diagnosed COVID (34.2 vs 99.3 days; Cox model HR+/-SE = 0.36 +/-0.24, p = 2.8×10^−5^, Figure 2B). As expected, the duration of the IgM response was significantly shorter than the IgG response (27.2 vs 54.3, Cox model HR+/-SE = 0.55 +/-0.09, p = 2.3×10^−10^, Figure 2C). Likewise, based on the antigen targets used by the two assays documenting time to sero-reversion, the humoral response to the nucleocapsid antigens (Syntron) was significantly shorter than the response to a combination of spike and nucleocapsid antigens (Innovita) (18.6 vs 49.8 days, Cox model HR+/-SE = 0.32 +/-0.11, p = 4.4×10^−16^, Figure 2D).

**Figure 2.**
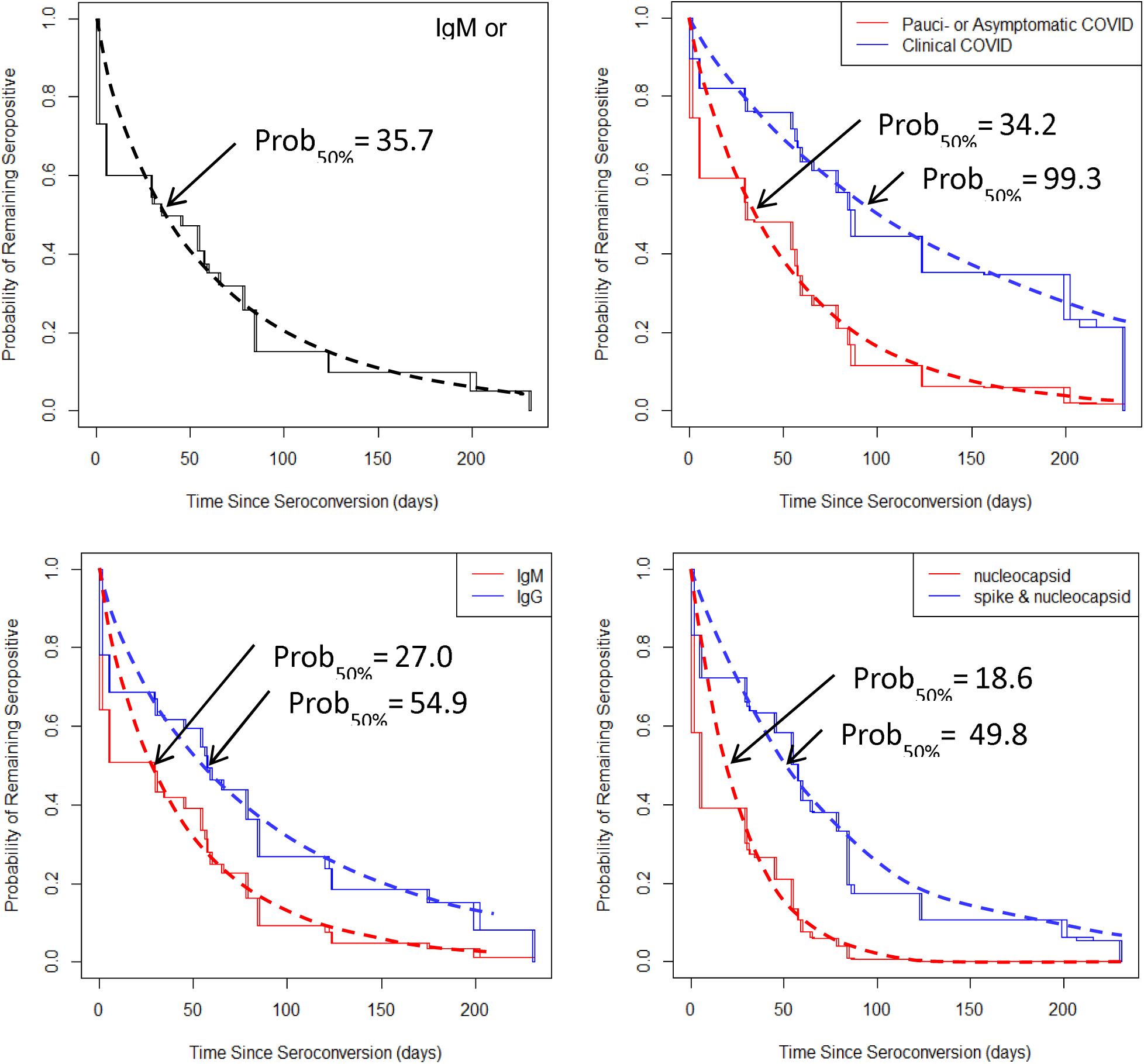
Semi-Parametric and Parametric (Weibull) Cox Proportional Hazard Models of Sero-reversion. Prob50% indicates the parametric estimate of time when 50% of the sero-positive cohort has become sero-negative. Panel A. Overall rate of sero-reversion for IgM or IgG. Panel B. Comparison of pauci- and asymptomatic vs. clinically-defined COVID cases. Panel C. Comparison of rates of IgM vs IgG sero-reversion. Panel D. Comparison of sero-reversion rates based on follow-up testing using the Syntron test targeting antibodies to nucleocapsid proteins and the Innovita test targeting a combination spike and nucleocapsid proteins.

## Discussion

In this study detectable antibody responses to SARS-CoV-2 in a largely pauci- or asymptomatic cohort were short-lived. Most cases sero-reverted in ∼30 days following documented sero-conversion. These data suggest that cross-sectional COVID-19 sero-surveillance studies have underestimated the population prevalence of prior infection.^2,5,17-24^ This observation has important implications for the epidemiology SARS-CoV-2. It suggests that community transmission of this pathogen may be even greater than currently presumed. By extension, estimates of hospitalization rate, infection fatality ratio and other measures of virulence, may also need to be revised downward. This in no way diminishes the magnitude of effect of this virus on public health. It simply highlights how pathogens causing morbidity and mortality in only a small percentage of cases can still pose a serious threat to public health when wide-spread community transmission occurs.

Not only does the short duration of elevated antibodies in minimally symptomatic cases make it difficult to discover them, it also raises a question about their long-term immunity. The answer to this question could have important implications for general public health interventions as well as the timing and targeting of population-wide interventions ^25^ – especially since the number of cases with an abbreviated humoral response is likely to be quite high. More data are needed on memory B- and T-cell generation and protection from re-infection in this large group of people with a clinically silent infection accompanied by a relatively brief humoral response.^26,27^

Recently, Lumley et.al reported results of longitudinal sero-surveillance in 452 healthcare workers following an initial positive SARS-CoV-2 serology result.^28^ Similar to the current study, they documented relatively rapid decay in IgG antibody titers over a period of several months, although a direct comparison of their estimated IgG half-life using a quantitative luminescent assay (85 days) and our estimate of IgG sero-positivity based on qualitative lateral flow assays (55 days) is not possible without a calibration of the lateral flow assays against the quantitative immunoassay. Importantly, in the UK study 61% of their participants recalled prior COVID-like symptoms and 21% had a positive SARS-CoV-2 PCR test as a result of symptomatic testing compared with the current study cohort which included predominately asymptomatic cases based on active daily symptom surveillance. In a separate study from the U.K. Ward et.al. reported declining rates of sero-positivity based on three distinct cross-sectional population-based surveys from June to September 2020.^29^ Although the sample size in this UK study was considerably larger than the current study, the absence of longitudinal data in the same subjects make it difficult to separate the effects of declining rates of detectable antibodies from changes in the background rate of new infections.

Much of the understanding of humoral responses to SARS-CoV-2 infection is based on small laboratory studies of people with clinically significant disease.^30-32^ Information on the kinetics of antibody responses in pauci- and completely asymptomatic cases are derived from an even smaller number of subjects with typically short follow-up and considerable variability in the definition of pauci- or asymptomatic cases.^8,9,33-36^ Nevertheless, these detailed laboratory studies consistently report that people with milder disease have a lower peak and a more rapid decline of SARS-CoV-2 specific IgG or IgM antibodies than more symptomatic cases. The current study, adds to this earlier work by providing considerable additional information on the large faction of cases in the population who have had little or no symptoms.

Ripperger et al ^4^ found that levels of IgG to the spike proteins (S2 and receptor binding domain) remained elevated much longer and more consistently than to the nucleocapsid proteins, including among volunteers with few or no symptoms. Our study provides evidence of a more durable response when focusing on IgG versus IgM, and on antibodies targeting spike and nucleocapsid versus exclusively nucleocapsid proteins. However, these effects are small relative to the overall picture of rapid sero-reversion observed in this cohort of mostly pauci- or asymptomatic SARS-CoV-2 infections.

The sample size in the current study allowed us to test for differences in time to sero-reversion as a function of age, sex, and race/ethnicity. Interestingly, among our mostly pauci- and asymptomatic cases none of these factors were related to time to sero-reversion. This is in contrast to associations between age and race/ethnicity and risk for symptomatic infection. ^37,38^ Understanding the factors that are associated with pauci- or asymptomatic infection with an abbreviated humoral response and clinically symptomatic disease with a more durable humoral response may provide novel insights about virology, immunology and clinical medicine with implications that extend beyond the current pandemic.

There are several limitations of our study. First, the sampling frame (two large healthcare system patient populations) and participants (volunteers) may reflect various biases including response bias that could influence rates of sero-conversion and sero-reversion in unknown directions. The preponderance of white participants and more female than male participants in the current study also raises questions about the generalizability of the results, although within the limits of statistical power afforded by the sample size, there were no clear difference in rates of sero-reversion by age, race/ethnicity or sex. The serology tests employed in this study were qualitative lateral flow assays. Although validated with convalescent samples in people with significantly elevated antibody titers, the calibration and comparison of these assays as a function of independently quantified antibody titers is not known. Some false positives and false negatives are likely. However, if we assume the test characteristics remained constant over time, and consider the consistency between the simple binomial rates and those estimated using interval censored repeated measures, we believe the observed change in rates of sero-positivity are reasonable estimates of the duration of detectable antibody responses in a population. The study design and contemporary factors related to the pandemic did not permit a regularly scheduled cadence of testing. Nevertheless, the data include a large number of tests with a continuous distribution over a wide period of time following an initial positive test allowing for good resolution in the estimates of time to sero-reversion. The COVID-19 Community Research Partnership has expanded to eight other medical centers to recruit additional participants for ongoing longitudinal surveillance. This will provide more data on antibody dynamics in primary infections and following vaccinations and support long-term clinical follow-up of asymptomatic cases to answer fundamentally important questions about how duration of initial antibody responses relate to the degree of subsequent protection from re-infection.

In summary, these data provide documentation of the duration of detectable antibody responses in a large number of mostly asymptomatic and minimally symptomatic cases of COVID-19. The short duration of the humoral response suggests that the true population prevalence of prior SARS-CoV-2 infection is likely significantly higher than presumed based on earlier sero-surveillance studies. The impact of the large number of cases with minimal symptoms and abbreviated antibody responses on population immunity remains to be determined.

Data Sharing Statement: At end of the study, the databases will be made publicly available in a de-identified manner according to CDC and applicable U.S. Federal policies. (https://covid19crp.bsc.gwu.edu/web/covid19crp/home).

## Writing Group

Wake Forest - David M Herrington MD, MHS, John Walton Sanders MD, MPH, Thomas F Wierzba PhD, Martha Alexander-Miller PhD, Mark Espeland PhD, Alain G. Bertoni MD, MS, Morgana Mongraw-Chaffin PhD, Allison Mathews PhD, Austin L. Seals MS, Iqra Munawar MS Atrium Health - Michael S. Runyon, MD, MPH, Lewis H. McCurdy, MD, Michael A. Gibbs, MD University of Maryland - Karen Kotloff, MD, DeAnna Friedman-Klabanoff, MD

MedStar - William Weintraub, MD

University of Mississippi – Adolfo Correa MD, PhD

George Washington Biostatistics Center - Diane Uschner PhD, Sharon Edelstein PhD, Michele Santacatterina PhD

## COVID-19 Community Research Partnership Study Group

### Wake Forest School of Medicine

John Walton Sanders MD, MPH, Thomas F Wierzba PhD, David Herrington MD, MHS, Mark A. Espeland PhD, Morgana Mongraw-Chaffin PhD, Alain Bertoni MD, Martha A. Alexander-Miller PhD, Allison Mathews PhD, Iqra Munawar MS, Austin Lyles Seals MS, Brian Ostasiewski, Christine Ann Pittman Ballard MPH

### George Washington Biostatistics Center

Diane Uschner PhD, Sharon L Edelstein ScM, Michele Santacatterina PhD, Greg Strylewicz PhD, Brian Burke MS, Mihili Gunaratne MPH, Meghan Turney MA, Shirley Qin Zhou MS

### Atrium Health

Michael S. Runyon MD, MPH, Lewis H. McCurdy MD, Michael A. Gibbs MD, Yhenneko Taylor PhD, Lydia Calamari MD, Hazel Tapp PhD, Amina Ahmed MD, Michael Brennan DDS, Lindsay Munn PhD, RN, Tim Hetherington MS, Lauren Lu, Connell Dunn, Melanie Hogg MS, CCRA, Andrea Price, Mariana Leonidas, Laura Staton, Kennisha Spencer MPH, Melinda Manning, Whitney Rossman MS, Frank Gohs MS, Anna Harris MPH, Bella Gutnik MS, Jennifer Priem PhD, MA

### MedStar

Kristen Miller DrPH, CPPS, William Weintraub MD, Chris Washington, Allison Moses, Sarahfaye Dolman, Julissa Zelaya-Portillo, John Erkus, Joseph Blumenthal, Romero Barrientos, Ronald E, Sonita Bennett, Shrenik Shah, Shrey Mathur, Christian Boxley, Paul Kolm, Long La, Cheng Zhang, Eva Hochberger, Ella Franklin, Deliya Wesley, Naheed Ahmed

### University of Maryland School of Medicine - Baltimore

Karen Kotloff MD, Wilbur Chen MD, MS, DeAnna Friedman-Klabanoff MD, Andrea Berry MD, Helen Powell, PhD

### Tulane University

Joseph Keating PhD, Patricia Kissinger PhD, Richard Oberhelman MD, John Schieffelin MD, Joshua Yukich PhD, Andrew “AJ” Beron MPH, Devin Hayes BS, Johanna Teigen MPH

### U of Mississippi

Adolfo Correa MD, PhD, Leandro Mena MD, MPH, Bhagyashri Navalkele MD, Yuan-I Min MD, Alexandra Castillo MPH, Lori Ward PhD, MS, Robert P. Santos MD, Courtney Gomillia MS-PHS, Pramod Anugu, Yan Gao MPH, Jason Green, Ramona Sandlin RHIA, Donald Moore MS, Lemichal Drake, Dorothy Horton RN

### WakeMed Health and Hospitals

William H. Lagarde MD, LaMonica Daniel BSCR

### New Hanover Regional Medical Center

Patrick D. Maguire MD, Lynette McFayden, RN

### Vidant Health

Thomas Gallaher, MD,Michael Zimmer, PhD,Shakira Henderson, PhD, DNP, MS, MPH, Danielle Oliver, Tina Dixon

### Campbell University

Robin King-Thiele DO, Terri S. Hamrick PhD, Chika Okafor MD, Regina B. Bray Brown MD, Pinoorma Vinod MD

### External Advisory Council

Helene Gayle MD/MPH, Chicago Community Trust (Chair), Ruth Berkelman MD, Emory, Kimberly Hanson MD, U of Utah, Scott Zeger PhD, Johns Hopkins, Cavan Reilly PhD, U. of Minn, Kathy Edwards MD, Vanderbilt,

The authors would also like to acknowledge the excellent programmatic and technical support provided by the dedicated staff at Vysnova Partners, Inc., Oracle Corporation, Scanwell Health, Inc. and Neoteryx.

### Role of Funding

This publication is supported by the CARES Act, of the U.S. Department of Health and Human Services (HHS) as part of a financial assistance award totaling $20,000,000. The contents are those of the author(s) and do not necessarily represent the official views of, nor an endorsement, by HHS, or the U.S. Government.

## Supplemental Figure

**Supplemental Figure 1.**
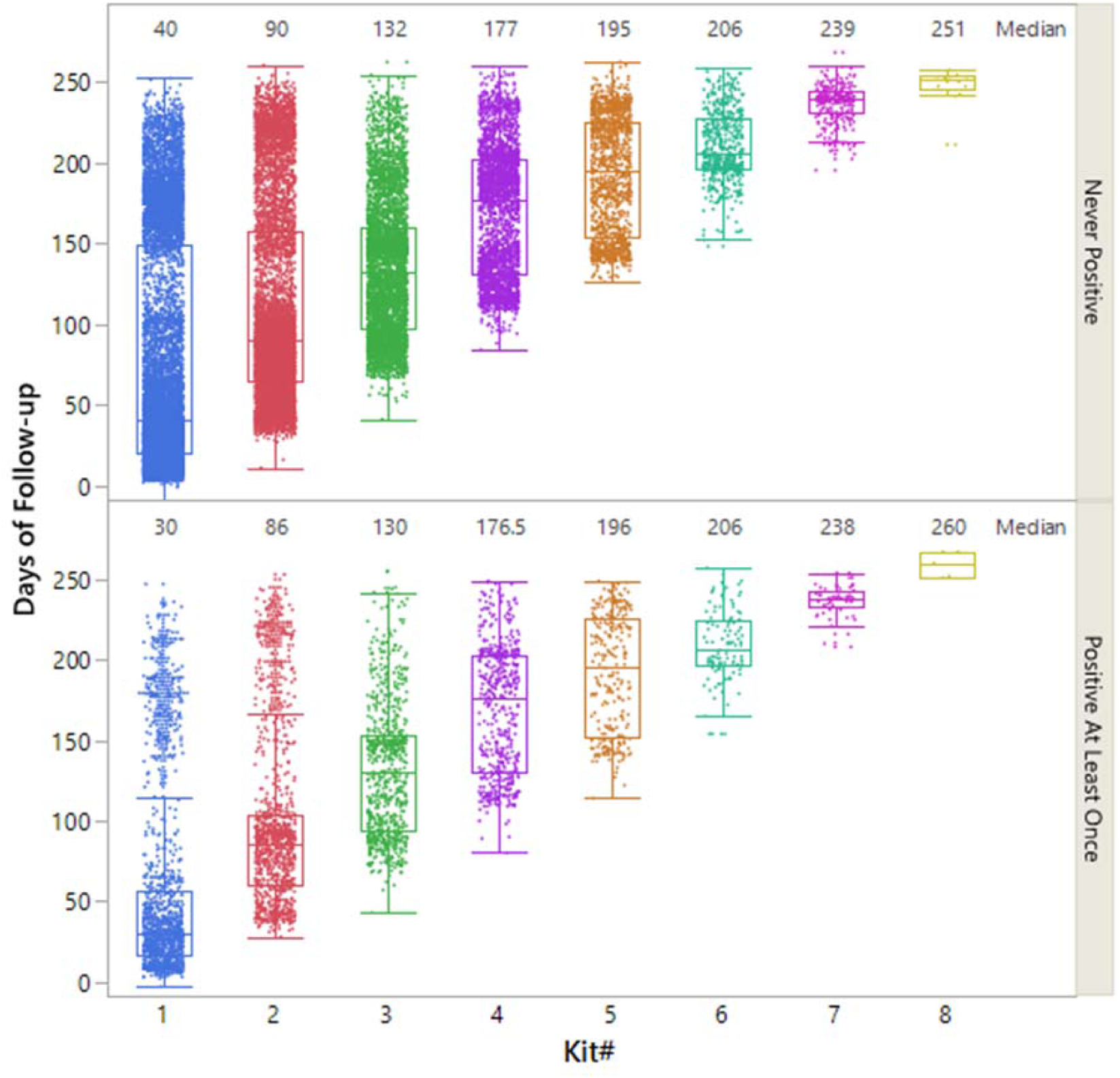
Distribution of longitudinal testing among participants that sero-converted vs those that remained negative during the period of follow-up.

